# Impact of the MyProstateScore (MPS) test on the clinical decision to undergo prostate biopsy: results from a contemporary academic practice

**DOI:** 10.1101/2020.03.17.20036384

**Authors:** Amir H. Lebastchi, Christopher M. Russell, Yashar S. Niknafs, Nicholas W. Eyrich, Zoey Chopra, Rachel Botbyl, Rana Kabeer, Takahiro Osawa, Javed Siddiqui, Rabia Siddiqui, Matthew S. Davenport, Rohit Mehra, Scott A. Tomlins, Lakshimi P. Kunju, Arul M. Chinnaiyan, John T. Wei, Jeffrey J. Tosoian, Todd M. Morgan

**Author notes:** **Corresponding authors:** Todd M. Morgan, MD, Associate Professor and Chief of Urologic Oncology, Department of Urology, The University of Michigan, 1500 E. Medical Center Dr., TC 387 / SPC 5330, Ann Arbor, Michigan 48109-5330, Jeffrey J. Tosoian, MD, MPH, Clinical Lecturer and Fellow, Urologic Oncology, Department of Urology, The University of Michigan, 1500 E. Medical Center Drive, TC 3875 SPC 5330, Ann Arbor, MI 48109. These authors contributed equally to this work (Amir H. Lebastchi, Christopher M. Russell). Co-senior authors (John T. Wei, Jeffrey J. Tosoian, Todd M. Morgan).

## Abstract

**Objective:** To evaluate the association of the MyProstateScore (MPS) urine test on the decision to undergo biopsy in men referred for prostate biopsy at a contemporary academic urology practice.

**Methods:** MPS testing was offered as an alternative to immediate biopsy in men referred to the University of Michigan for prostate biopsy from October 2013 through October 2016. The primary endpoint was the decision to perform biopsy. The proportion of patients who underwent biopsy was calculated across MPS quintiles and compared to predicted risk scores from the Prostate Cancer Prevention Trial risk calculator (PCPTrc). Analyses were performed in the overall referral population and the intended-use population (PSA 3-10 ng/ml or PSA <3 ng/ml and abnormal DRE) and stratified by the use of multiparametric magnetic resonance imaging (mpMRI). The associations of PCPTrc, MPS, and mpMRI with the decision to undergo biopsy were explored in a multivariable logistic regression model.

**Results:** Of 248 patients, 134 (54%) proceeded to prostate biopsy. Clinical variables, PSA, and PCPTrc score did not significantly differ based on the decision to undergo biopsy, while MPS was significantly higher in biopsied patients (median 29 vs. 14, p<0.001). The use of biopsy was strongly associated with MPS, with biopsy rates of 26%, 38%, 58%, 90%, and 85% in the first through fifth MPS quintiles, respectively (p<0.001). By contrast, biopsy rates were 51%, 47%, 52%, 69%, and 52% by increasing PCPTrc score quintile (p=0.3). The association of MPS with biopsy persisted upon stratification by use of mpMRI. On multivariable analysis, MPS was strongly associated with the decision to undergo biopsy when modeled as both a continuous variable (odds ratio [OR] 1.05 per 1 unit increase, 95% confidence interval [CI] 1.04-1.08; <0.001) and binary variable (OR 7.76, 95% CI 4.14-14.5; p<0.001). These findings were consistent in the overall and intended-use populations.

**Conclusion:** In a cohort of patients who underwent clinical MPS testing as an alternative to immediate prostate biopsy, 46% were able to avoid biopsy and increasing MPS was strongly associated with biopsy rates. These findings were robust to the use of mpMRI and consistent across pertinent subpopulations. Overall, these data suggest that the MPS assay may substantially reduce the number of men undergoing prostate needle biopsy.

## Introduction

Early detection of prostate cancer continues to pose a substantial clinical challenge. While over one million prostate biopsies are performed annually due to elevated serum prostate-specific antigen (PSA), fewer than 50% of biopsies will detect cancer and even fewer will detect clinically-significant cancer.^1–4^ The poor specificity of serum PSA for cancer results in widespread use of biopsy and unnecessary detection of low-risk cancers – two sources of significant morbidity.^5,6^ The limitations of PSA testing have driven the development of novel biomarkers aimed at avoiding unnecessary biopsies (i.e. negative biopsies and those detecting low-grade cancer),^7–9^ while accurately detecting higher-risk cancers more likely to benefit from early detection.^10^

The MyProstateScore (MPS) test (previously termed Mi-Prostate Score, or MiPS) combines serum PSA with urinary PCA3 and TMPRSS2:ERG (T2:ERG) to better inform the risk of high-grade (Grade Group [GG] ≥2) cancer.^11^ Urinary PCA3 has been shown to increase the sensitivity and specificity of testing for GG≥2 cancer, and its use is approved by the US Food and Drug Administration to facilitate decision-making in men with previous negative biopsies.^1,12,13^ The T2:ERG gene fusion is present in 50% of prostate cancer foci and 70% of men with prostate cancer, and it exhibits >99% specificity for cancer in tissue.^14–18^ Several previous studies have demonstrated superior diagnostic accuracy of PCA3 and T2:ERG relative to PSA,^7,18,19^ and their combined use in MPS has significantly outperformed multivariable models incorporating PSA plus clinical data, such as the Prostate Cancer Prevention Trial risk calculator (PCPTrc).^11^ For example, in one recent multicenter study, the combined use of PCA3 and T2:ERG would have yielded a 42% reduction of unnecessary biopsies, while preserving high sensitivity (≥93%) for significant cancers.^20^

While the validity of MPS has been demonstrated, the impact of MPS testing on clinical decision-making has not been reported. We sought to characterize the association of MPS testing with the decision to undergo subsequent prostate biopsy in men referred for biopsy at a single academic institution.

## Methods

### Study cohort

From October 2013 through November 2016, following Institutional Review board approval, MPS was ordered as an alternative to immediate biopsy in 248 men referred to the University of Michigan for prostate biopsy (i.e. elevated PSA, concerning PSA kinetics, and/or abnormal digital rectal examination [DRE]). Use of MPS was based on provider discretion, and the final study cohort included all men who underwent MPS testing as part of the diagnostic workup. Baseline clinical variables included age, race, first-degree family history of prostate cancer, DRE findings, and history of previous biopsy. For patients who underwent multiparametric magnetic resonance imaging (mpMRI), findings were recorded based on the prostate imaging reporting and data system (PI-RADS) v2.^21^

The decision to pursue prostate biopsy was made by the physician and patient through shared decision-making. Prostate needle biopsies were performed under transrectal ultrasound (TRUS) guidance using a standard 12-core sextant template, and mpMRI-based regions of interest were targeted in most cases (i.e. 9 of 12 patients (75%) with PI-RADS 3 lesions and 20 of 22 patients (91%) with PI-RADS ≥4 lesions). All biopsy specimens were reviewed by dedicated genitourinary pathologists in accordance with the 2014 International Society of Urological Pathology Consensus Conference.^22^

### Urine MPS generation

Urine processing for PCA3 and T2:ERG scores was performed on post-DRE whole urine specimens using transcription-mediated amplification (TMA) assays, as previously described.^7,19,23^ Details of the PCA3 and T2:ERG (Progensa) assays are provided in the Supplement*. MPS values were calculated using the previously published, locked regression models including only serum PSA, urinary PCA3 score, and urinary T2:ERG score.^11^ MPS values were reported on a continuous scale from 0 (very unlikely to detect GG≥2 cancer on prostate biopsy) to 100 (very likely to detect GG≥2 cancer on prostate biopsy).

### Statistical analysis

To assess the impact of MPS on clinical behavior, the primary study endpoint was the decision to perform prostate biopsy. As a measure of clinical validity, detection of high-grade cancer (GG≥2) was a secondary endpoint in patients that underwent biopsy. Baseline characteristics were assessed in patients who did and did not undergo biopsy, and comparisons were made using the Mann-Whitney test for continuous variables and chi-squared test for proportions. The risk of high-grade cancer was calculated for all patients based on the PCPTrc version 2.0, which includes age, race, serum PSA, DRE results, family history of prostate cancer, and history of previous negative biopsy.^24^ Because validated MPS thresholds were not established at the time of the study and MPS results were reported along a risk continuum, the proportion of patients that underwent prostate biopsy was considered by quintile of MPS and PCPTrc score. Findings were unchanged in binary and quartile-based sensitivity analyses (Supplement*).

Analysis was performed in both the (i) overall biopsy referral population and (ii) an intended-use population of men with PSA 3-10 ng/ml or PSA < 3 ng/ml and abnormal DRE, as indicated by the National Comprehensive Cancer Network (NCCN) clinical practice guidelines.^25^ Multivariable logistic regression was used to explore the association of clinical variables (based on the PCPTrc and as individual variables), MPS, and mpMRI on the decision to perform biopsy. Primary analyses used a six-month follow-up interval from MPS testing to identify associated biopsies.^26^ Findings were similar in sensitivity analyses based on three- and twelve-month follow-up intervals (Supplement*). Analyses were performed using R version 3.6.1.

## Results

### Study population

During the study period, 248 men presented to the University of Michigan Department of Urology with suspicion of prostate cancer and underwent MPS testing as an alternative to immediate prostate biopsy. Of these patients, 134 (54%) subsequently underwent prostate biopsy. As shown in **Table 1**, age, race, family history, DRE status, and history of previous biopsy were not significantly different in patients who did versus did not undergo biopsy.

**Table 1.** Cohort characteristics by decision to pursue biopsy

|  | Overall (n=248) | No biopsy (n=114) | Biopsy (n=134) | P-value |
| --- | --- | --- | --- | --- |
| Age | 66 (61-70) | 65 (61-69) | 66 (61-70) | 0.98 |
| Black race | 16 (6.5%) | 9 (7.9%) | 7 (5.2%) | 0.4 |
| Family history | 56 (23%) | 27 (24%) | 29 (22%) | 0.7 |
| Abnormal DRE | 36 (15%) | 16 (14%) | 20 (15%) | 0.8 |
| Previous biopsy | 158 (64%) | 72 (63%) | 86 (64%) | 0.9 |
| PSA (ng/ml) | 7.1 (4.8-10.1) | 6.2 (4.4-10.2) | 7.5 (5.3-9.9) | 0.08 |
| PCPTrc | 10% (7-15) | 9% (7-15) | 10% (7-15) | 0.2 |
| MPS | 24 (13-41) | 14 (9-25) | 29 (22-48) | <0.001 |

PCPTrc = Prostate Cancer Prevention Trial risk calculator for high-grade cancer, version 2.0. MPS = MyProstateScore.

Similarly, serum PSA (median 6.2 ng/ml vs. 7.5 ng/ml, p=0.08) and high-grade PCPTrc score (10 vs. 9, p=0.2) did not differ by subsequent use of biopsy. MPS was significantly higher in men who proceeded to biopsy compared to those who did not (median 29 vs. 14, p<0.001).

### Primary outcome: Decision to perform biopsy

In the absence of validated threshold values, the association of MPS and PCPTrc with the decision to undergo biopsy was assessed by quintile values. As illustrated in **Figure 1A**, the proportion of patients undergoing prostate biopsy was strongly associated with increasing MPS values. Specifically, only 13 men (26%) with a first quintile MPS (0-11) and 19 (38%) with a second quintile MPS (11-17) proceeded to biopsy, as compared to 36 (58%), 40 (90%), and 37 (85%) patients in the third, fourth, and fifth MPS quintiles, respectively (p<0.001). By contrast, the decision to pursue biopsy was not associated with PCPTrc score quintile, as the proportion of men proceeding to biopsy in the first through fifth PCPTrc score quintiles were 51%, 47%, 52%, 69%, and 52% (**Figure 1B**, p=0.3). These relationships were unchanged in the analysis limited to the intended-use population (**Figure 2**).

**Figure 1.**
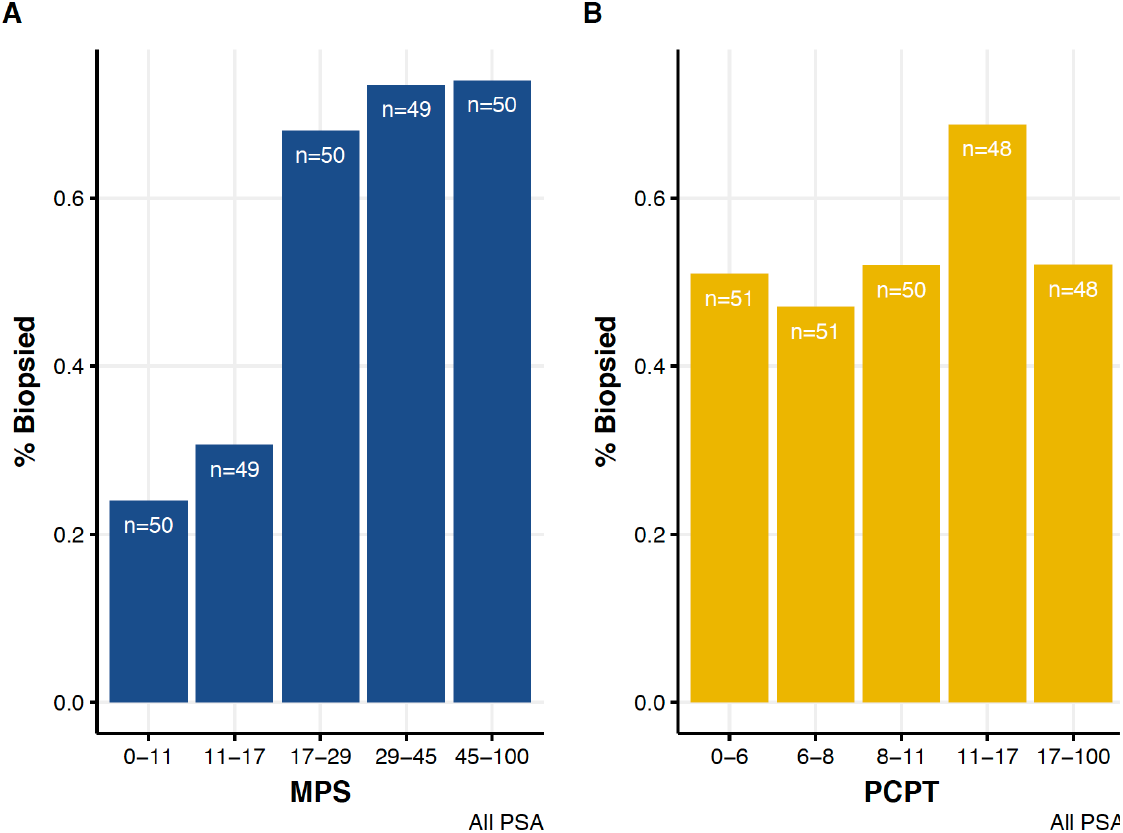
Proportion of patients who underwent prostate biopsy by quintile scores of A) MPS and B) PCPTrc in the overall population (n=248).

**Figure 2.**
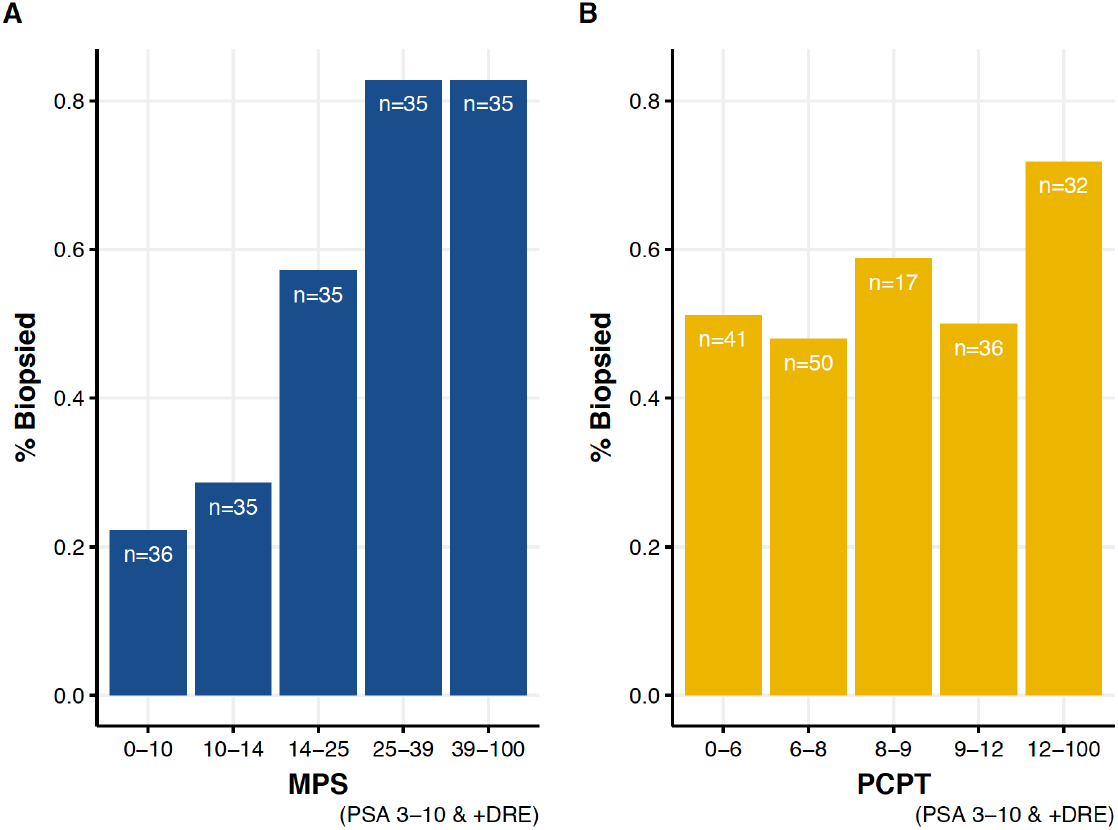
Proportion of patients who underwent biopsy by quintile scores of A) MPS and B) PCPTrc in the intended-use population (i.e. PSA 3-10 ng/ml or PSA < 3 ng/ml and abnormal DRE) (n=172).

### Primary outcome: In context of mpMRI

There were 87 patients (35%) who also underwent mpMRI prior to clinical decision-making, and 55 (62%) of these men proceeded to biopsy (vs. 49% of those without mpMRI, p=0.03). Consistent with contemporary practice, PIRADS≥3 lesions were significantly more common in mpMRI patients who proceeded to biopsy (62% vs. 31% in those without biopsy, p=0.006). Notably, MPS was significantly associated with the decision to undergo biopsy regardless of mpMRI use. Among patients who did not undergo mpMRI, median MPS was 30 (21-47) in men who underwent biopsy and 14 (10-25) in men who did not (p<0.001). In the MRI population, median MPS values were 29 (22-52) and 13 (6-25) in those who did and did not undergo biopsy, respectively (p<0.001). **Figure 3** illustrates MPS and PI-RADS scores by biopsy status (i.e. no biopsy, negative/GG1, GG≥2).

**Figure 3.**
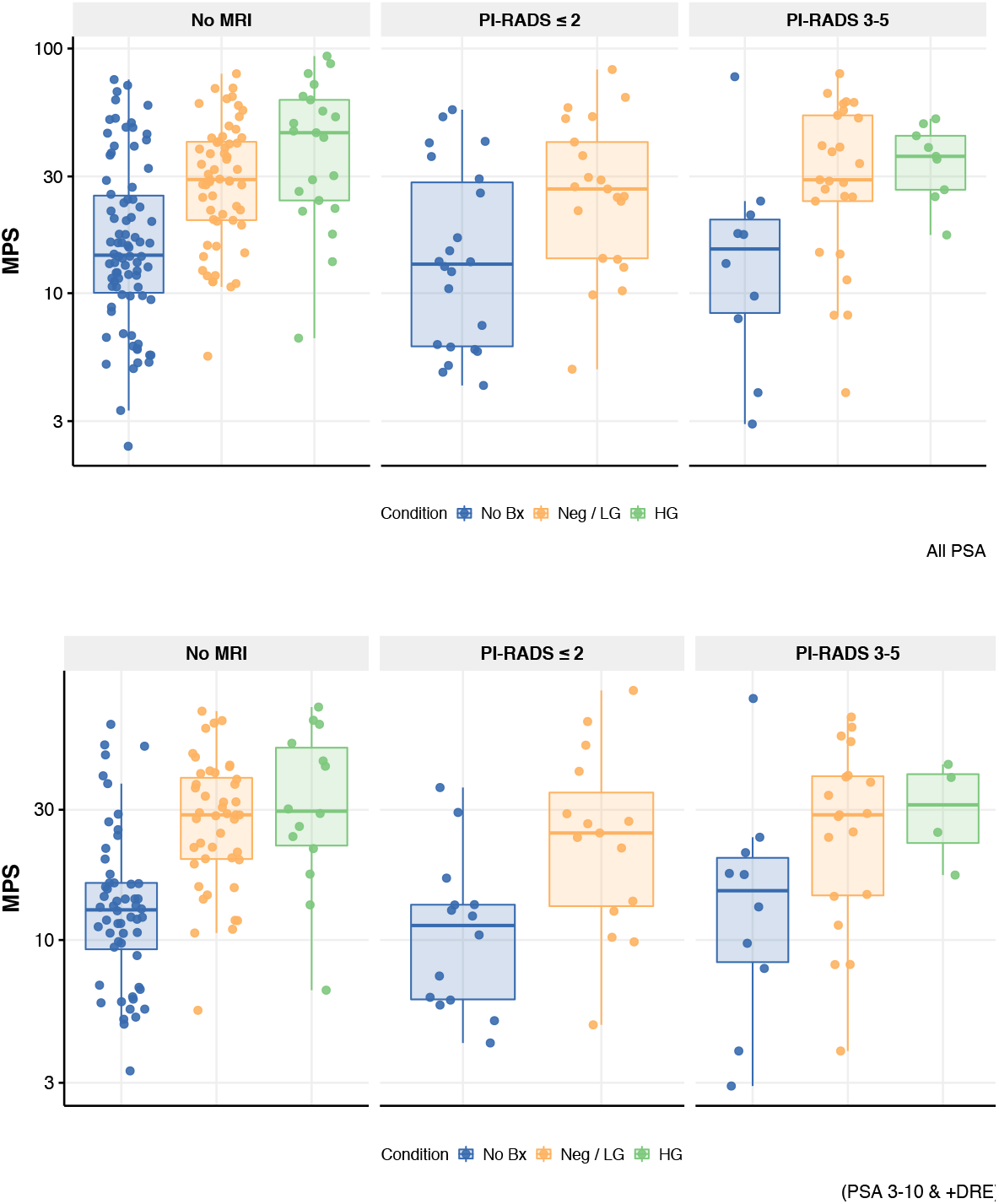
Box and whisker plots of MPS values by PI-RADS score and biopsy status. Blue = did not undergo biopsy; yellow = biopsy was negative or revealed Grade Group 1 cancer; green = biopsy revealed Grade Group 2 cancer. Data are presented for A) the overall population (n=248) and B) the intended-use population (i.e. PSA 3-10 ng/ml or PSA < 3 ng/ml and abnormal DRE) (n=172).

### Primary outcome: Multivariable models for biopsy decision-making

We explored the relationship of MPS, mpMRI, and clinical variables (i.e. PCPTrc) with the primary outcome (i.e. decision to undergo biopsy) in multivariable logistic regression models. As displayed in **Table 2**, higher MPS values were strongly associated with the decision to undergo biopsy. This association was present when MPS was parametrized as a continuous variable (OR 1.05 per 1 unit MPS increase, 1.04-1.08, p<0.001), by MPS quintiles (OR 2.05, 1.60-2.61, p<0.001), and dichotomized relative to the median value (OR 6.81, 3.65-12.7, p<0.001). As compared to men that did not undergo mpMRI, positive mpMRI (i.e. PI-RADS 3-5) was also associated with the decision for biopsy, while negative mpMRI was not (**Table 2**). In the subpopulation that underwent both MPS testing and mpMRI (n=87), increasing MPS was strongly and significantly associated with use of biopsy in all models, while PI-RADS score was not (**Table 3**). These findings were unchanged when clinical variables were considered independently rather than in the combined PCPTrc (Supplement*).

**Table 2.** Multivariable logistic regression models for the association of PCPTrc, mpMRI, and MPS with the decision to perform biopsy in the overall population (n=248)

|  | Model 1: MPS Continuous | Model 2: MPS Quintiles | Model 3: Binary MPS |
| --- | --- | --- | --- |
|  | OR (95% CI; p-value) | OR (95% CI; p-value) | OR (95% CI; p-value) |
| PCPTrc (per 1 unit inc.) | 0.96 (0.92-0.99; 0.02) | 0.97 (0.94-1.00; 0.02) | 0.98 (0.95-1.01; 0.2) |
| mpMRI results (vs. no MRI) |  |  |  |
| PI-RADS 1-2 | 1.33 (0.63-2.81; 0.5) | 1.33 (0.62-2.84; 0.5) | 1.07 (0.51-2.27; 0.9) |
| PI-RADS 3-5 | 3.34 (1.47-7.58; 0.004) | 3.27 (1.42-7.51; 0.005) | 3.18 (1.38-7.32; 0.007) |
| MPS (per 1 unit inc.) | 1.05 (1.04-1.08; <0.001) | - | - |
| MPS quintile (per 1 unit inc.) | - | 2.05 (1.60-2.61; <0.001) | - |
| MPS ≥ median (vs. <24) | - | - | 6.81 (3.65-12.7; <0.001) |

PCPTrc = Prostate Cancer Prevention Trial risk calculator for high-grade cancer, version 2.0. MPS = MyProstateScore.

**Table 3.**
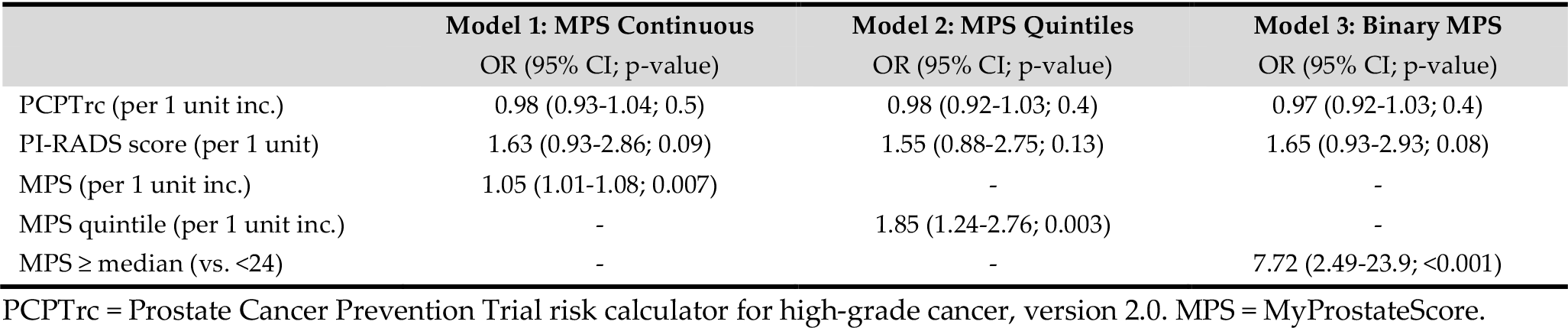
Multivariable logistic regression models for the association of PCPTrc, PI-RADS score, and MPS with the decision to perform biopsy in patients that underwent mpMRI (n=87)

|  | Model 1: MPS Continuous | Model 2: MPS Quintiles | Model 3: Binary MPS |
| --- | --- | --- | --- |
|  | OR (95% CI; p-value) | OR (95% CI; p-value) | OR (95% CI; p-value) |
| PCPTrc (per 1 unit inc.) | 0.98 (0.93-1.04; 0.5) | 0.98 (0.92-1.03; 0.4) | 0.97 (0.92-1.03; 0.4) |
| PI-RADS score (per 1 unit) | 1.63 (0.93-2.86; 0.09) | 1.55 (0.88-2.75; 0.13) | 1.65 (0.93-2.93; 0.08) |
| MPS (per 1 unit inc.) | 1.05 (1.01-1.08; 0.007) | - | - |
| MPS quintile (per 1 unit inc.) | - | 1.85 (1.24-2.76; 0.003) | - |
| MPS ≥ median (vs. <24) | - | - | 7.72 (2.49-23.9; <0.001) |

PCPTrc = Prostate Cancer Prevention Trial risk calculator for high-grade cancer, version 2.0. MPS = MyProstateScore.

### Secondary outcome: Biopsy pathology

Among the 134 patients who underwent prostate biopsy, 80 men (60%) had a negative biopsy, while 24 men (18%) had GG1 disease, 14 (10%) had GG2, 10 (7%) had GG3, and 6 (4%) had GG 4-5. **Figure 4A** illustrates the distribution of MPS values in men who did not undergo biopsy (n=114), men who underwent a negative/GG1 biopsy (n=104), and those who underwent a GG≥2 biopsy (n=30). Notably, MPS was significantly higher in men with GG≥2 cancer compared to those with negative biopsy or GG1 disease (median 39 vs. 29, p=0.02). Similar relationships were observed in the subgroup of patients with PSA 3-10 ng/ml (**Figure 4B**), although the difference in MPS between the negative/GG1 and GG≥2 biopsy groups was minimal (median 29 vs. 30, pairwise p=0.3).

**Figure 4.**
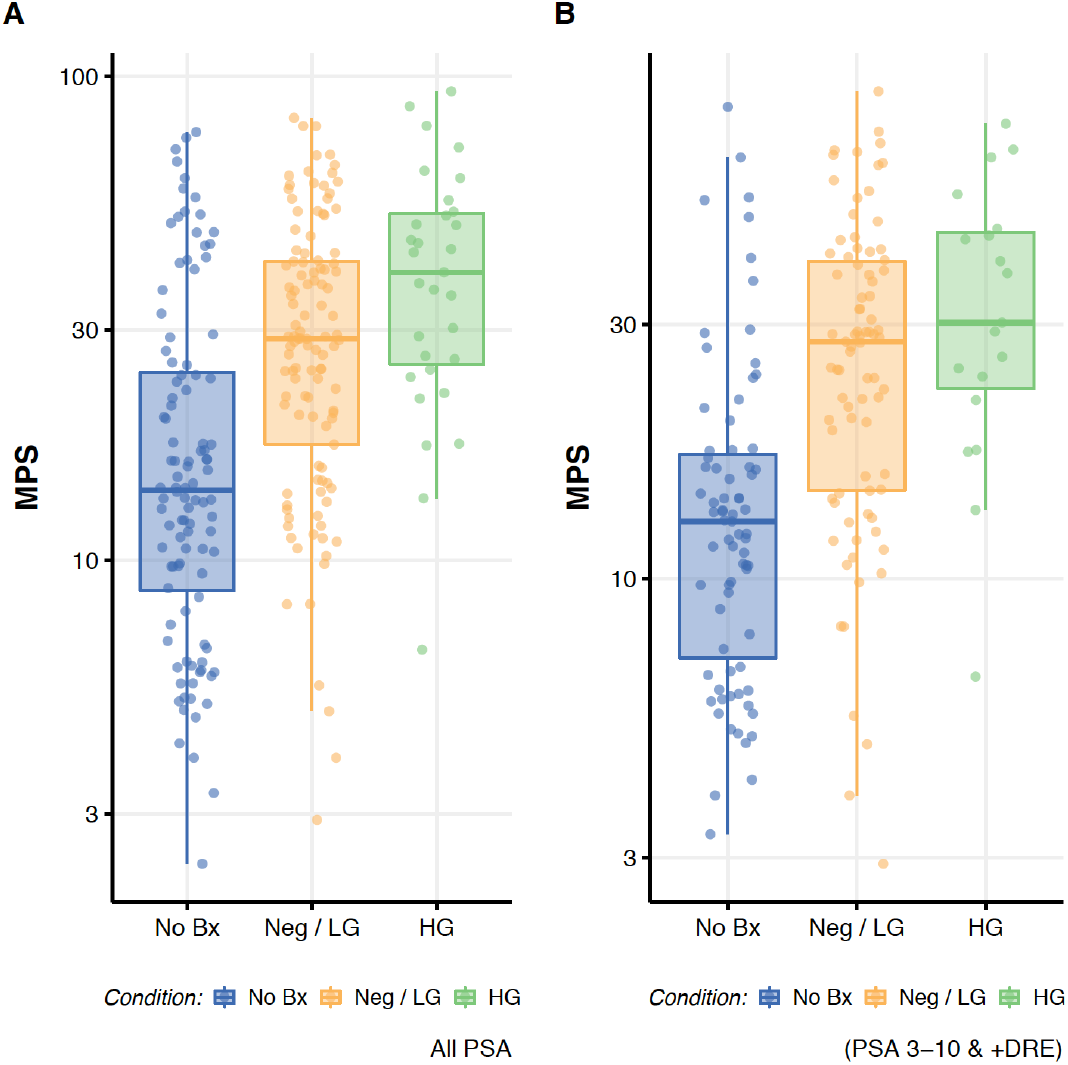
Box and scatter plot of MPS values stratified by choice to pursue biopsy and biopsy results in the A) overall population (n=248) and B) PSA 3-10 ng/ml population (n=172).

## Discussion

We assessed the impact of MPS on the decision to perform prostate biopsy in 248 consecutive patients who underwent MPS testing as an alternative to immediate biopsy. In total, 46% of the tested cohort was able to avoid biopsy during six month follow-up. Notably, MPS values were strongly associated with the decision to perform subsequent biopsy, with only 26% of men in the lowest MPS quintile and 38% of men in the second MPS quintile undergoing biopsy, as compared to 76% in the remaining population. By contrast, scores from the PCPT risk calculator for GG≥2 cancer, which combines serum PSA with age, race, serum PSA, DRE results, family history of prostate cancer, and history of previous negative biopsy, did not appear to impact biopsy rates. Importantly, the association of MPS with the decision to perform biopsy was present in men who did and did not undergo mpMRI. Ultimately, these findings suggest that the information provided by MPS impacted clinical decision-making and appeared to reduce the use of biopsy.

The clinical validity of MPS and its component markers have been demonstrated in a number of previous reports.^11,19,23,27,28^ These studies have found that urinary PCA3 and T2:ERG scores improve the predictive ability of pre-biopsy testing relative to current diagnostic models such as the PCPTrc. In a validation cohort of 1225 men presenting for prostate biopsy, Tomlins et al. assessed the discriminatory performance of models for GG≥2 cancer using the area under the receiver operating characteristic curve (AUC). The observed AUC was 0.65 for PSA alone, 0.71 for PSA plus clinical variables (i.e. PCPTrc), and 0.77 for MPS.^11^ Notably, the addition of clinical variables to the MPS model did not meaningfully improve AUC (0.772 vs. 0.779). Thus, the validated, clinically available MPS test does not depend on clinical variables and patient history, which can be subjectively measured and unreliably recorded. Use of an objectively measured non-invasive test obtained during routine care, such as MPS, is likely to provide substantial improvement over current practice.

The National Comprehensive Cancer Network (NCCN) early detection guidelines for prostate cancer suggest that physicians consider the use of biomarkers or mpMRI in patients indicated for biopsy (i.e. PSA 3-10 ng/ml or PSA < 3 ng/ml and abnormal DRE).^25^ As such, we described our observations in the overall population referred for biopsy and specifically among the intended-use population defined by the NCCN. We found that the association of MPS with the decision to undergo biopsy was consistent across these populations. The consistency of these findings across subgroups is in line with published validation data, in which MPS-based models better predicted GG≥2 cancer than PSA- and PCPTrc-based models in all subgroups (28/28 = 100%) stratified by serum PSA (≤ 3, 3-10, >10 ng/ml), DRE findings, and history of previous biopsy.^11^ Combined, these data suggest a potential role for MPS in better defining the risk of high-grade cancer prior more invasive testing (i.e. biopsy or mpMRI) in men traditionally subjected to immediate prostate biopsy.

As described, mpMRI was performed in 87 study participants, and MPS was strongly associated with the decision to perform biopsy after stratification by use of mpMRI. In regression models exploring the relationship of MPS and mpMRI with biopsy, MPS was strongly and significantly associated with biopsy across all models. PI-RADS score was also associated with use of biopsy, although this relationship did not meet conventional levels of statistical significance. Both MPS and mpMRI demonstrated high validity, as there was only one case of GG≥2 cancer in men with MPS ≤10 (i.e., first-quintile values; Figure 2) and none in men with negative mpMRI (PI-RADS ≤2). Given the limited number of patients who underwent both tests (n=87), our primary aim was to account for the impact of mpMRI on the relationship between MPS and biopsy, rather than to identify an optimal combined approach to using MPS and mpMRI. Nonetheless, considering both (i) the practical limitations of mpMRI (e.g. subjective interpretation, high inter-reader variability, high relative cost and resource burden)^29,30^ and (ii) the very high negative predictive value of MPS (98%) on recent validation,^31^ MPS could be useful as a first-line test to rule-out high-grade cancer prior to more invasive testing.

There are limitations of the current study. First, these data reflect a study period in which MPS thresholds for clinical use were not proposed or validated, and MPS results were provided across a range from 0 to 100. Thus, there was no objective measure of a “negative” or “low-risk” MPS, and the decision to pursue biopsy was ultimately based on discretion. Thus, the association of MPS with the primary outcome was based on numerical categorization (i.e. quintiles). Nonetheless, the strength of association observed across MPS quintiles as compared to traditional variables indicates that MPS test results impacted clinical decision-making. Second, this was a single-arm study with no comparison cohort without MPS testing. We therefore assessed the association between MPS and biopsy relative to clinical information (PCPTrc). Third, while measures of the proportion biopsied imply a baseline 100% biopsy rate in the absence of MPS or mpMRI, it is likely that other considerations could have reduced use of biopsy in the absence of MPS. Still, the lack of association between clinical data and biopsy questions the appropriateness of such an approach. Also, findings from this single academic practice may not represent the population at-large, supporting the need for additional assessments of utility. Finally, as noted, the pathology of patients who did not undergo biopsy is unknown, thus the current study should not be considered a robust assessment of clinical validity. As this was not the primary objective, it is nonetheless notable that the limited assessment of validity performed herein was consistent with existing data

## Conclusions

In a cohort of patients referred for prostate biopsy, initial use of MPS testing was associated with the avoidance of biopsy in 46% of men. Notably, MPS results were strongly associated with the decision to proceed to biopsy, regardless of the use of other tools such as the PCPTrc and mpMRI. Combined with the body of data supporting clinical validity of MPS, these findings suggest utility of MPS testing to rule out the need for more invasive assessments such as mpMRI and biopsy. The current analysis merits controlled, prospective assessment of the impact of MPS testing on clinical decision making.

## Data Availability

Data associated with article can be provided on request. Please contact the corresponding author.

## Acknowledgements

JJT is supported by the National Institutes of Health/National Cancer Institute Advanced Training in Urologic Oncology Grant (T32/CA180984). His research is funded in part by an award from the SPORE Career Enhancement Program (CA186786).

AMC is a Howard Hughes Medical Institute Investigator and an American Cancer Society Research Professor.

This work was supported by the Prostate Cancer Foundation, Early Detection Research Network (UO1 CA214170), NCI Prostate SPORE (P50 CA186786), and an NCI Outstanding Investigator Award (R35CA231996).

TMM is supported by the A. Alfred Taubman Medical Research Institute.

## Potential Conflicts of Interest

JJT, YSN, and AMC are co-founders and have equity in Lynx Dx, which has licensed the urine biomarkers mentioned in this study from Hologic and the University of Michigan. JJT and YSN have leadership roles in Lynx Dx. The University of Michigan has been issued a patent on ETS gene fusions in prostate cancer on which AMC and SAT are co-inventors. The diagnostic field of use has been licensed to Lynx Dx. SAT serves as CMO of Strata Oncology which was not involved in this study. Lynx Dx or Strata Oncology did not fund the conduct of this study.

## Notes

### Clinical Trial

This was an institutional review board-approved retrospective analysis evaluating the impact of a clinically-available test on clinical decision making. Thus it was not a prospectively registered trial.

